# Toward Standards for Donor-Level Metadata Reporting in Human Tissue Research: Perspectives from Tissue Researchers

**DOI:** 10.64898/2026.07.24.26358862

**Authors:** Rachel Liu-Galvin, Kathy Tran, Mishal Khan, Michael T. Eadon, Pinaki Sarder, Yulia A. Levites Strekalova

**Affiliations:** Department of Health Services Research, Management and Policy, College of Public Health and Health Professions, University of Florida, Gainesville, FL, USA; Department of Economics, College of Liberal Arts and Sciences, University of Florida, Gainesville, FL, USA; Divisions of Nephrology and Clinical Pharmacology, Indiana University School of Medicine, Indianapolis, IN, USA; Department of Medicine (Quantitative Health), College of Medicine, University of Florida, Gainesville, FL, USA

**Keywords:** Tissue Donors, Biomedical Research, Reproducibility of Results, Confidentiality, Ethics, Research

## Abstract

**Introduction:** No widely adopted guidelines exist for collecting and reporting donor-level metadata in tissue-based research, limiting interpretability, reproducibility, and potentially introducing bias. This study aimed to inform ethical and appropriate metadata practices.

**Methods:** Semi-structured interviews were conducted with 16 investigators from the Human BioMolecular Atlas Program. Thematic analysis using inductively derived codes identified metadata elements and perspectives on their collection and reporting.

**Results:** Participants identified 80 metadata variables across six domains: demographic, sociodemographic, medical history, personally identifying information, cause of death, and tissue/organ data. Most supported routine collection and reporting of demographics and medical history, whereas views on cause of death and sociodemographic data were mixed.

**Conclusion:** We recommend routinely collecting and reporting demographics and medical history, while restricting cause of death and sociodemographic variables to situations with explicit consent or justification. These findings provide initial evidence to inform ethical donor metadata guidelines, with further stakeholder engagement and consensus-building needed.

## INTRODUCTION

Human tissue research relies on donor samples, yet there remains no consensus on which donor information should be ethically and scientifically reported.^1,2^ While tissue processing protocols are described in detail, donor-level metadata such as age, sex, medical history, and socioeconomic indicators are inconsistently captured.^3^ These metadata may provide important contextual information about exposures affecting epigenetic regulation.^4^ Furthermore, this gap between human characteristics and cell-level findings creates a broken link in the data lifecycle, limiting reproducibility, introducing potential biases, and reducing the interpretability of results.^3,5^ These challenges underscore the need for standardized guidelines for reporting donor-level metadata in studies using human tissue samples.

While cell-level methods such as single-cell RNA sequencing, spatial transcriptomics, and spatial epigenomics, have significantly advanced our understanding of human biology, less attention has been given to capturing human characteristics known to influence cellular function and physiology through epigenetics.^4,6–8^ Contextual information such as demographics, medical and drug histories, and geographic location reveal exposures that may influence gene expression.^4^ These mechanisms underscore the importance of collecting donor-level metadata in tissue-based research, while balancing scientific value with privacy and identifiability concerns.^2^

Biospecimen studies often underrepresent minority groups and healthy controls, compromising external validity.^9^ To produce accurate, generalizable, and equitable results, donor samples should reflect the U.S. population. Non-representative data introduce bias, perpetuate health disparities, and undermine the validity of models trained on these data, leading to inaccurate or systematically biased predictions.^10^ The lack of donor-level metadata for tissue samples is a recognized barrier to reproducibility and clinical translation.^11^ Promoting inclusive and transparent reporting is therefore critical. Despite this, there are no widely adopted guidelines defining the specific demographic, clinical, and contextual variables that constitute a complete donor profile for tissue-based research, the necessary granularity for each variable, and how to ensure consistent reporting.^12^ While guidelines exist for randomized trials, these are not suited for cellular-level tissue research, due to their treatment of social variables, such as race, as proxies for biological variables, such as genotype, without clear justification.^13^

To our knowledge, this study is the first to address this gap by obtaining researchers’ perspectives to inform guidelines for the appropriate and ethical collection and reporting of donor-level metadata in tissue-based research. Using interviews with investigators from the National Institutes of Health (NIH)-funded Human BioMolecular Atlas Program (HuBMAP), we identified consensus and points of contention on which donor-level metadata should be collected or reported, establishing a baseline from which to initiate a broader community effort to standardize the human side of tissue-based research.

## METHODS

### Setting and Participants

This qualitative elicitation study recruited participants from the NIH-funded HuBMAP consortium, an international initiative focused on mapping the human body at cellular resolution.^18^ HuBMAP investigators work with human tissue samples and associated data, making them well-positioned to provide insight into which donor-level metadata should be collected for tissue-based research.

Participants were purposively sampled from active HuBMAP members in leadership or senior technical roles (e.g., principal investigators, professors, research scientists, and laboratory directors) to capture perspectives across key workstreams, including tissue procurement, experimental biology, data generation, and bioinformatics. Participants represented nine institutions across academia, healthcare, and biotechnology sectors, providing diverse perspectives on human tissue donation.

Participants were contacted via email up to three times. Recruitment was guided by information power considerations to ensure sufficient depth for informing donor data collection recommendations.^19^

### Interview Data Collection

Fifteen interviews were conducted with sixteen participants (including one interview that involved two participants) via secure video conferencing between July and November 2024. Participants received a standardized explanation of the project and provided verbal consent prior to participation.

Interviews were conducted individually by trained qualitative researchers with no prior relationship to participants to minimize social desirability bias. A semi-structured guide (Appendix C) explored perspectives on representation in research, donor tissue heterogeneity, and implications for data quality and scientific advancement. Interviews lasted between 6 and 31 minutes (mean: 17 minutes) and were audio-recorded and transcribed verbatim using Microsoft 365 Transcribe, with manual verification for accuracy.

### Data Analysis

Transcripts were cleaned (e.g., spelling, abbreviations, filler words) prior to analysis. The lead author developed a preliminary codebook through iterative review of the transcripts, which was refined in collaboration with co-authors. Thematic analysis was conducted using inductively derived codes to capture the full breadth of metadata elements mentioned by participants. The final codebook (Appendix E) categorized variables into demographic information, sociodemographic information, medical history, personally identifying information, cause of death, and tissue or organ data (e.g., sample quality, viability, and other sample features). For each variable, participants’ views were classified into three categories: (1) collect and report, (2) collect but not report, or (3) neither collect nor report. A presence/absence-based coding approach was used, with each variable coded once per interview to prioritize breadth of perspectives.

Variables were classified as ‘collect and report’ unless participants explicitly recommended otherwise; routine practices were interpreted as endorsements unless objections were noted. This interpretative approach was considered appropriate given the open-ended nature of the interview questions and is consistent with established qualitative research practices, in which inference and interpretation are integral to the analytic process.^20^

Two investigators independently coded all transcripts, followed by a joint reconciliation phase to achieve consensus. To enhance interpretability, 80 variables were synthesized into broader categories (e.g., ‘laboratory test results’) as shown in Appendix E. Analyses were conducted using R version 4.4.1 and RStudio.

## RESULTS

We conducted 15 interviews with 16 participants (including one two-participant interview) and asked them for their opinions on the data that should be collected from tissue donors. Our thematic analysis identified 80 donor-level metadata variables, consolidated into 25 variable types. Variable types were further grouped into five categories: 1) medical history, 2) demographic information, 3) sociodemographic information, 4) cause of death, 5) tissue or organ data (e.g., sample quality, viability, or other sample features), and 6) personally identifying information. Participants recommended whether each variable be 1) collected and reported, 2) collected but not reported, or 3) neither collected nor reported. Appendix A provides full variable mapping by transcript; aggregate frequencies are summarized in Appendix B. Consolidated frequencies of categories stratified by recommendation type are presented in Table 1, which lists variable categories in order of “collect and report” recommendations.

Figure 1 depicts the distribution of recommendations across categories, and Appendix C presents the distribution of recommendations by variable type.

Most variables within the medical history category were recommended for full collection and reporting. Physiological measurements (e.g., BMI) received the most mentions, followed by laboratory test results; diagnosed conditions and general medical histories were also frequently cited. However, a few participants suggested withholding certain measurements or test results from publications. Some participants also suggested collecting lifestyle data, such as diet, physical activity, and substance use beyond tobacco and alcohol, to provide insight into cellular-level health effects.^4^ However, obtaining and standardizing this information from tissue donors is challenging, as it would likely depend on secondary sources such as medical records or next-of-kin reports, or on instruments such as 24-hour recall interviews, food-frequency questionnaires, or wearable devices, which are impractical in the context of tissue donation.

Demographics were universally recommended for collection and reporting. Of these, race and age were most frequently mentioned, followed by ethnicity, sex, and gender. A minority noted some demographics, particularly race, should only be published with donor consent. Participants emphasized that race and ethnicity help ensure donor representativeness, although one interviewee noted the limitation of race being a “man-made construct”, consistent with ongoing discussions in biomedical research.^13^

Views on sociodemographic context were mixed; with collection and reporting recommended by most participants, while some suggested this information, particularly socioeconomic status and geographic location, should be collected but not reported, or omitted altogether. Several participants recommended collecting socioeconomic status, an important non-clinical determinant of health,^4^ although no standard instrument exists for capturing this information from donors. One possible tool is the Area Deprivation Index (ADI),^14^ a neighborhood-level measure of socioeconomic disadvantage, but its reliance on zip code data raises concerns of identification; this may necessitate explicit donor or next-of-kin consent and could be challenging to implement in practice.

For tissue samples obtained from deceased donors, few participants recommended recording and cause and mechanism of death. Among those who did, most recommended collecting and reporting it, while a minority recommended collection only, omitting its inclusion in publications.

Anatomical site of tissue collection and distance of healthy and diseased tissue came up as a variable category that we name tissue provenance. Participants recommended recording and reporting tissue or organ sample features such as sample quality and anatomical characteristics. Molecular data (e.g., genomic data) were also mentioned, although a minority of participants suggested they should be collected but not reported. Because these variables relate to specimen-level characteristics, they were captured during coding but not included in the final table of recommendations, which focuses on donor-level metadata.

Personally identifying information was discussed frequently, with most participants recommending this data be neither collected nor reported. A few participants suggested certain information, e.g., zip code and social security number, be collected but not reported. Only one interview recommended collecting and reporting a socioeconomic disadvantage derived from zip code (Appendices A and B).

**Table 1.**
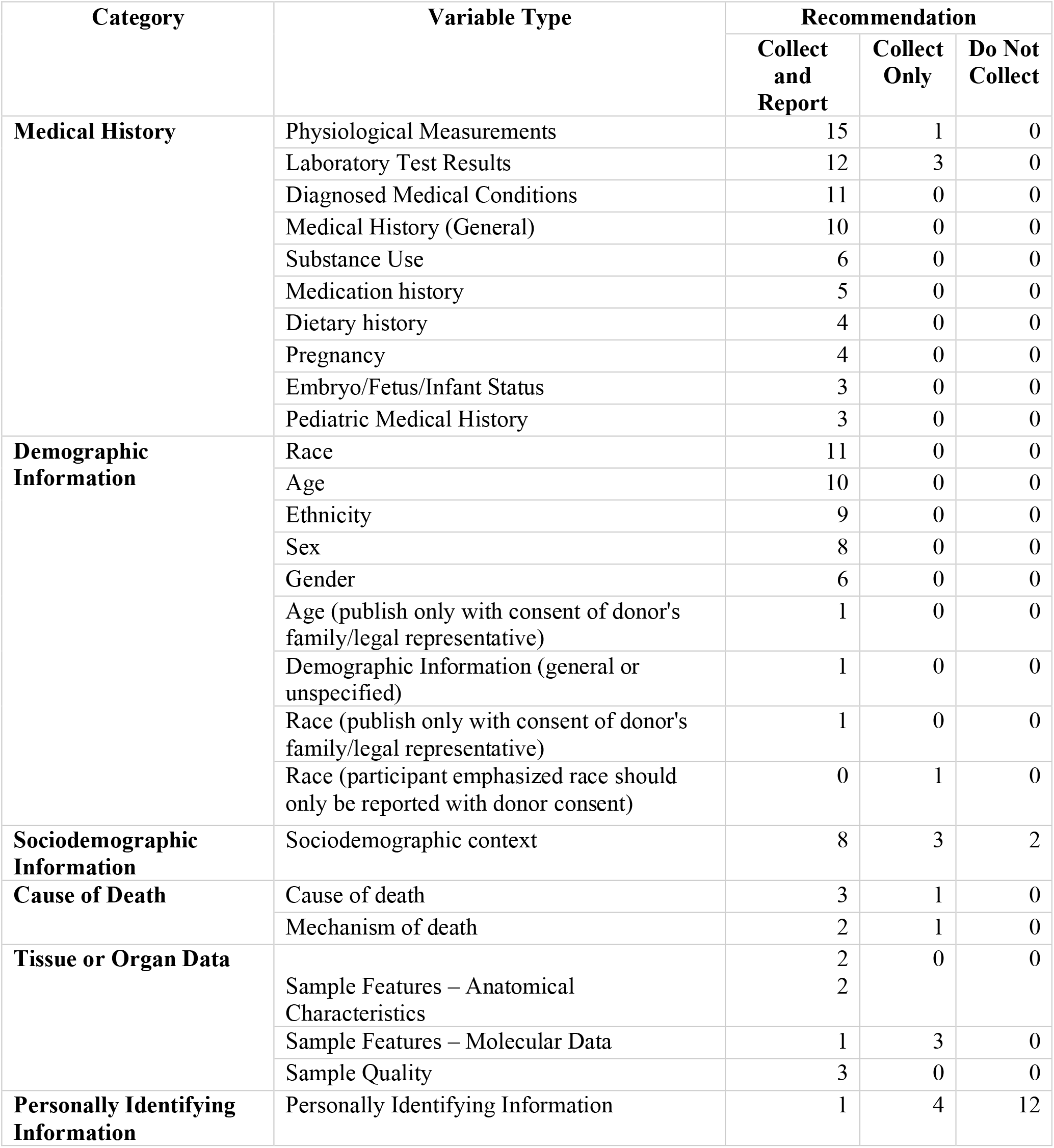
Frequency of Collection and Reporting Recommendations from HuBMAP Investigators.

Based on our findings, we propose the following recommendations for the collection and reporting of donor-level metadata in tissue-based research. Items with strong consensus were endorsed by most participants and may serve as clear priorities for inclusion in future guidelines. Items with mixed perspectives highlight areas where further research and consensus assessment are needed. These tiered recommendations are summarized in Table 2.

**Table 2.** Participant-Informed Recommendations for Donor-Level Metadata Collection and Reporting.

| **Recommendation Category** | **Metadata Elements** | **Summary of Participant Perspectives** |
| --- | --- | --- |
| **Collect and Report** | Demographic information: age, race, ethnicity, sex, and gender | Strong consensus for both collection and reporting; minority recommended reporting age or race only with donor or family consent |
|  | Medical history: physiological measurements (e.g., height, weight, BMI, laboratory results, blood pressure), laboratory test results, diagnosed medical conditions, substance use, medication history, dietary history, pregnancy-related medical history (if applicable), medical history related to the embryo, fetus, or infant (if applicable), and pediatric medical history (if applicable) | Strong consensus for both collection and reporting |
| **Collect but Not Routinely Report** | Cause and mechanism of death | Mixed perspectives; we recommend collecting this data internally, and reporting only with explicit donor/family consent or when a strong scientific justification outweighs re-identification risk |
|  | Sociodemographic context | Mixed perspectives; we recommend collecting this data internally, and reporting only with explicit donor/family consent or when a strong scientific justification outweighs re-identification risk |
| **Do Not Collect** | Personally identifying information, e.g., name, address, zip code, telephone numbers, social security number | Strong consensus against collection due to substantial privacy concerns and high risk of donor re-identification |

## DISCUSSION

This study provides a novel, stakeholder-informed perspective on the importance of capturing donor-level metadata in tissue-based research. Since data use and ethical requirements vary across institutions and jurisdictions, this study does not seek to standardize these factors, but rather to document investigators’ views to inform guidelines. A key strength of this study was the integration of insights from investigators engaged in tissue-based research, supporting the development of future stakeholder-informed guidelines that balance scientific utility with participant privacy and practical feasibility.

Interviews were conducted by two researchers and coded independently by two others to reduce bias, although, as the coders were not present during interviews, some nuances of participant responses may not have been captured. Additionally, the semi-structured interview format, involving open-ended questions, gives rise to inherent subjectivity of interpretation.

To our knowledge, this is the first study to systematically explore which donor-level metadata should be collected and reported in tissue-based research. Existing reporting standards such as CONSORT, STROBE, or the National Cancer Institute (NCI)’s Best Practices for Biospecimen Resources are tailored to clinical, trials, epidemiological studies, and oncology, respectively, and do not specify which donor-level metadata should be published in tissue-based studies.^15–17^ This leaves a clear gap that our findings aim to address. Our recommendations align with the NCI’s minimal clinical data set for annotating biospecimen resources, with several variables in common including demographics, medical history, and lifestyle factors. While several specimen-level variables were also mentioned by participants, these were not included in our final table of recommendations because our aim was to identify donor-level metadata that could contextualize cell-level findings.

Our findings show strong consensus on reporting demographics, medical history, and tissue or organ sample quality; in contrast, cause and mechanism of death and sociodemographic context elicited mixed views. These findings provide an initial evidence base to inform the development of guidelines for collecting and reporting donor-level metadata in tissue-based research. Future studies should gather input from a wider variety of stakeholders, such as tissue and organ procurement organizations, regulatory bodies, e.g., the United Network for Organ Sharing (UNOS), and donor advocacy groups, to capture diverse perspectives. Areas that elicited mixed views warrant further clarification through consensus-building approaches such as Delphi expert panels. Developing and adopting clear, ethical donor metadata guidelines is essential for advancing accurate, equitable, reproducible, and generalizable tissue-based research that translates into improved population health.

## ACKNOWLEDGEMENTS

We would like to acknowledge Nicole Nash for her contributions to data collection.

## DATA AVAILABILITY STATEMENT

The data for this study are available from the authors upon reasonable request.

## ETHICAL APPROVAL

This study was reviewed and approved by the Institutional Review Board (IRB) at the University of Florida ET00041509.

